# Buprenorphine Initiation by a Street Medicine Program is Associated with Reduced Opioid-Related Emergency Department Utilization Among People Experiencing Homelessness

**DOI:** 10.1101/2025.09.17.25336012

**Authors:** Cecilia R. Wallace, Jasmine King, Timothy Nichols, Elizabeth A. Abrams, Shreya Shaw, Nicole K. Hoeflinger, Vidya Mullangi, Brittany E. Sandoval, Josh Matzke, Emily Kauffman, Irene Mynatt, Melissa R. Pelc, Ryan Cantzler

## Abstract

**Objectives:** People experiencing homelessness are at increased risk for addiction, with substance use both a cause and response to homelessness. Street Medicine (SM) programs provide decentralized, low-barrier care to people experiencing homelessness and have begun to offer medication for opioid use disorder (MOUD). Literature on MOUD initiation via SM is nascent. This study investigated the impact of SM-initiated MOUD on ED utilization among people experiencing homelessness in a mid-sized US city.

**Methods:** We completed a retrospective medical record review of patients initiated on buprenorphine in 2023 by the SM program. Age, gender, prescription refill status, and emergency department (ED) utilization were recorded. Outcome measures were the number of overdose-related, opioid use disorder (OUD)-related, and all-cause ED visits in the 12 months before versus 12 months after buprenorphine initiation.

**Results:** In the year prior to buprenorphine initiation, 115 patients had 221 cumulative ED visits, 79 of which were OUD-related. In the year following initiation, cumulative ED visits declined to 191, of which 44 were related to OUD (p<0.05). Overdose-related ED visits decreased from 26 to 13. In terms of prescription renewal, 44% of subjects renewed, 13% via SM and 31% via another provider.

**Conclusions:** Buprenorphine initiation by a SM program was associated with a significant decline in OUD-related ED visits and insignificant reductions in overdose-related and total ED visits. These conclusions support SM-initiated buprenorphine as a strategy for reducing opioid-related ED utilization while encouraging sustained engagement in care. SM programs represent promising avenues to initiate buprenorphine and reduce OUD-related morbidity in the unhoused population.

## 1. Introduction

A record-high 653,104 people experienced homelessness in the United States on a single night in January 2023, with more than 40% living unsheltered.^1,2^ This growing national crisis has been increasing over the past two decades.^3^

People experiencing homelessness (PEH) face substantially higher rates of premature mortality compared to the general population, primarily due to the higher prevalence of medical comorbidities, mental illness, and substance use disorders.^4–6^ Notably, drug overdose— especially opioids—accounts for 25-50% of deaths within this population, a trend that has been intensifying over the past decade.^7^ The risk of overdose related death in this vulnerable population is augmented by fragmented healthcare services and limited access to overdose prevention tools, such as naloxone.^7,8^ Moreover, due to the lack of consistent primary care, homeless patients are far more likely to seek emergency care, utilizing the emergency department (ED) at more than three times the rate of the general population.^9^ These disruptions in continuity of care further the poor health outcomes among PEH.

The growing homelessness epidemic underscores the urgency of improving health outcomes. Specifically, targeting opioid overdose-related morbidity and mortality could significantly improve the health of people experiencing homelessness and lower the rates of premature mortality seen in this population. It is crucial to implement comprehensive strategies to provide substance use education and treatment to this medically and socially vulnerable population.

One plausible intervention is to increase access to medication for opioid use disorder (MOUD) among these communities. MOUD treats symptoms of withdrawal, reduces opioid cravings, and facilitates long-term recovery.^10^ It has demonstrated strong efficacy in reducing rates of overdose, and is proven to improve quality of life overall among people who use drugs seeking recovery.^11–15^ Access to MOUD is limited for the general population, with nearly 87% of individuals with opioid use disorder (OUD) not receiving this treatment.^16^ For PEH, this challenge is compounded by the significant barriers homelessness imposes on healthcare access. Factors such as lack of transportation, financial difficulties, and challenges in obtaining medications make initiating and maintaining adherence to MOUD particularly difficult for PEH. Moreover, efforts to decentralize MOUD, such as offering MOUD at mental health treatment facilities and shelters are limited.^17,18^ Given the prevalence of opioid use and overdose rates among PEH, developing effective solutions to increase access to MOUD is critical.

A model that improves treatment accessibility among this population is through employing mobile medical units, such as street medicine (SM) programs, to prescribe MOUD. Street medicine seeks to decentralize medical care and offer treatment and social support directly within the spaces that PEH are living in, whether that be tent encampments, on the streets, or within shelters.^19^ SM programs offer a variety of services, from wellness checks to providing immediate treatment for acute medical concerns. Several urban SM programs have successfully implemented low-barrier access to MOUD, leading to high patient retention rates in treatment programs and reduced opioid use.^20^ Although there have been success stories with this model, the literature on the long-term impact of SM programs on ED utilization and overdose rates is limited. The purpose of our study is to understand the impact of MOUD initiation on ED utilization among patients served by a private, urban, midwestern SM program.

## 2. Methods

### 2.1 Study Design & Setting

This retrospective cohort study was performed using data from an urban community hospital’s SM program. The SM team has over 4,000 patient encounters annually includes physicians, nurses, bilingual case workers, and a community paramedic. All visits are tracked using a comprehensive electronic medical record system. This study was approved by the hospital’s Institutional Review Board (IRB #240627-3).

### 2.2 Participants

A team member blind to the study objectives generated a dataset of electronic medical records of all patients prescribed buprenorphine by the SM program from January 1 to December 31, 2023. Inclusion criteria included adult patients (aged 18 and above) whose prescription was initiated by the SM program. Patients whose buprenorphine was continued but not initiated by the SM program were excluded.

### 2.3 Data Collection

Data abstraction from medical records was completed by four abstractors using a structured template. Abstractors were trained by two physicians from the SM program. Medical record abstractors met regularly to discuss progress and monitor for discrepancies. To ensure data quality and accuracy, a SM physician verified the data collected from each medical record after the initial round of abstraction. There were no missing data.

### 2.4 Outcome Measures

Patient characteristics recorded were age, sex, and prescription renewal status. The primary outcome of interest was the number of ED visits in the 12 months before and after buprenorphine start date, subcategorized as visits due to (a) all-cause, (b) overdose, opioid use, or withdrawal (e.g., abscess, cellulitis, endocarditis, notated as “OUD-related”) and (c) overdose-only. Reason for visit was determined by reviewing ED chief complaints. If it was unclear whether the chief complaint was OUD-related, then ED notes were reviewed in detail.

### 2.5 Analysis

We conducted two complementary analyses to evaluate changes in ED utilization before and after buprenorphine initiation.

To evaluate cohort-level changes in ED utilization rates we calculated cumulative cohort visits before and after buprenorphine initiation. We fit negative binomial regression models to cumulative visits before and after buprenorphine, determining incident rate ratios (IRR) representing the relative change in ED visit rates after buprenorphine initiation.

To assess individual-level changes in ED utilization, we performed Wilcoxon signed-rank tests on all-cause, OUD-related, and overdose-specific ED visits. This non-parametric, paired test allowed us to evaluate individuals’ ED use change.

Next, to examine whether the intervention’s effectiveness varied across different patient characteristics, we conducted subgroup analyses on demographic variables: sex, age (younger [<36 years old] versus older [36 and over]), and prescription renewal status (any renewal versus no renewal). Prescription renewal status was defined as collecting another buprenorphine prescription (prescribed by SM or another provider) as reported by statewide Prescription Drug Monitoring Program (PDMP) data; or not collecting another buprenorphine prescription within the study period. For each patient, we calculated an ED utilization change score by subtracting the number of ED visits in the 12 months before the intervention from the number of visits in the 12 months after. We examined all-cause, OUD-related, and overdose-specific ED utilization change by subgroup.

We used the non-parametric Wilcoxon rank-sum test (aka Mann-Whitney U test) to compare change scores between subgroups. For each comparison, we calculated descriptive statistics including median changes and interquartile ranges to characterize the magnitude and variability of changes within each subgroup. Statistical significance was set at p < 0.05 for all comparisons. Analyses were completed using R version 4.4.2 within RStudio.

## 3. Results

### 3.1 Patient Characteristics

The SM program prescribed buprenorphine to 134 patients during the study period. Of these, 19 were excluded due to buprenorphine initiation outside of the SM program, leaving 115 in analysis. Patient characteristics are shown in Table 1. Within the 12 months after buprenorphine initiation, 51 patients collected a renewed prescription, with 16 seeking continued MOUD care from the SM program and 35 establishing with another provider. Of the 64 patients who did not collect a renewed prescription within the study window, 6 patients were known to engage in long-term MOUD or residential treatment after the 12-month period following buprenorphine initiation.

**Table 1.** Patient characteristics.

| Table 1. Patient characteristics |  |  |
| --- | --- | --- |
|  |  | Overall (N=115) |
| Age range (years) |  |  |
|  | 18-25 | 5 (4.3%) |
|  | 26-35 | 35 (30.4%) |
|  | 36-45 | 45 (39.1%) |
|  | 46-55 | 26 (22.6%) |
|  | 55+ | 4 (3.5%) |
| Gender |  |  |
|  | Male | 68 (59.1%) |
|  | Female | 47 (40.9%) |

| Prescription renewal source |  |  |
| --- | --- | --- |
|  | Street med | 16 (13.9%) |
|  | Other | 35 (30.4%) |
|  | No renewal | 64 (55.7%) |

### 3.2 ED Utilization

In the 12 months prior to buprenorphine initiation by the SM program, this cohort of patients accounted for 221 all-cause ED visits. In the 12 months following initiation, this declined to 191 all-cause ED visits (Table 2, Figure 1). While all-cause visits decreased by 13.6%, this change was not significant (IRR = 0.864, 95% CI: 0.576-1.297, p = 0.481). Individual-level analysis showed a trend toward reduced utilization that did not reach statistical significance (p = 0.075), with the median number of visits per person decreasing from 1 (IQR: 0-3) to 0 (IQR: 0-2).

**Table 2.**
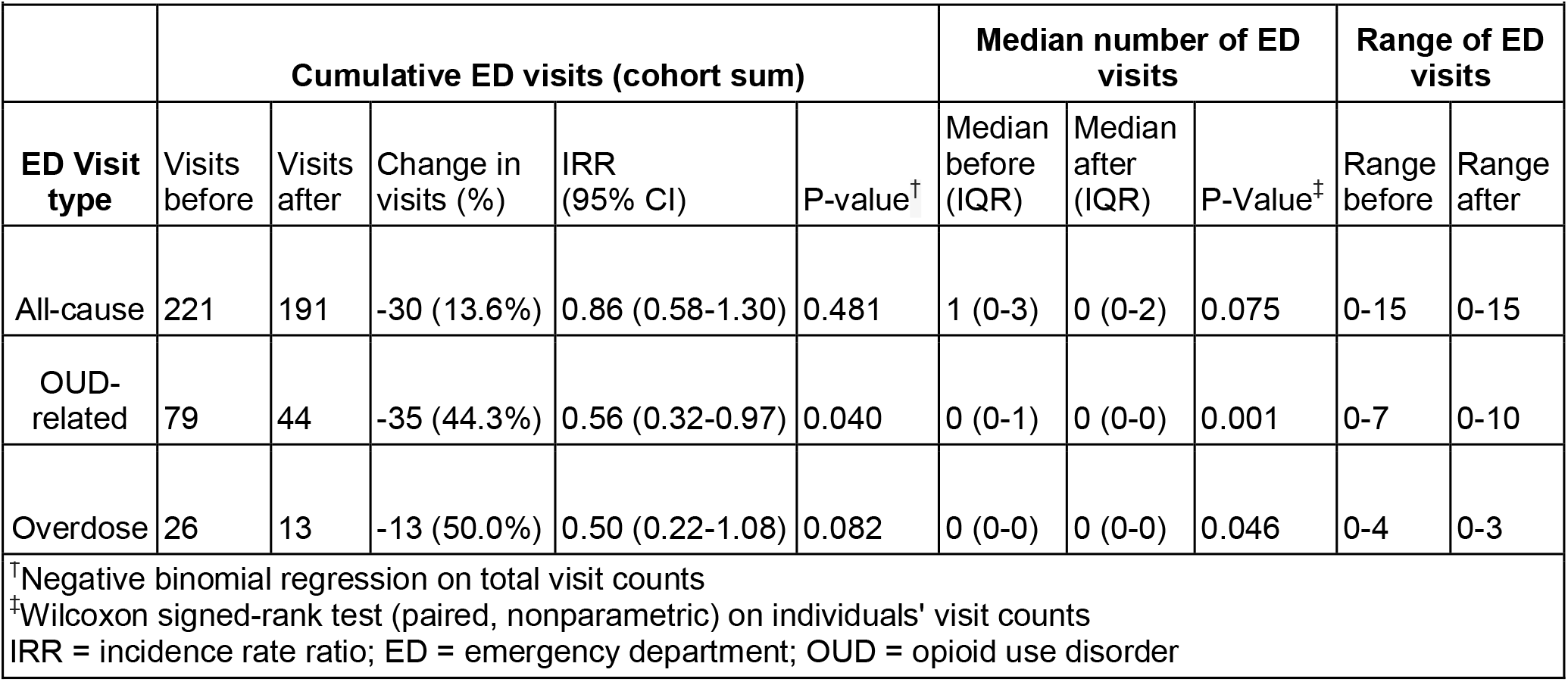
Summary of ED visits before and after buprenorphine initiation.

**Figure 1.**
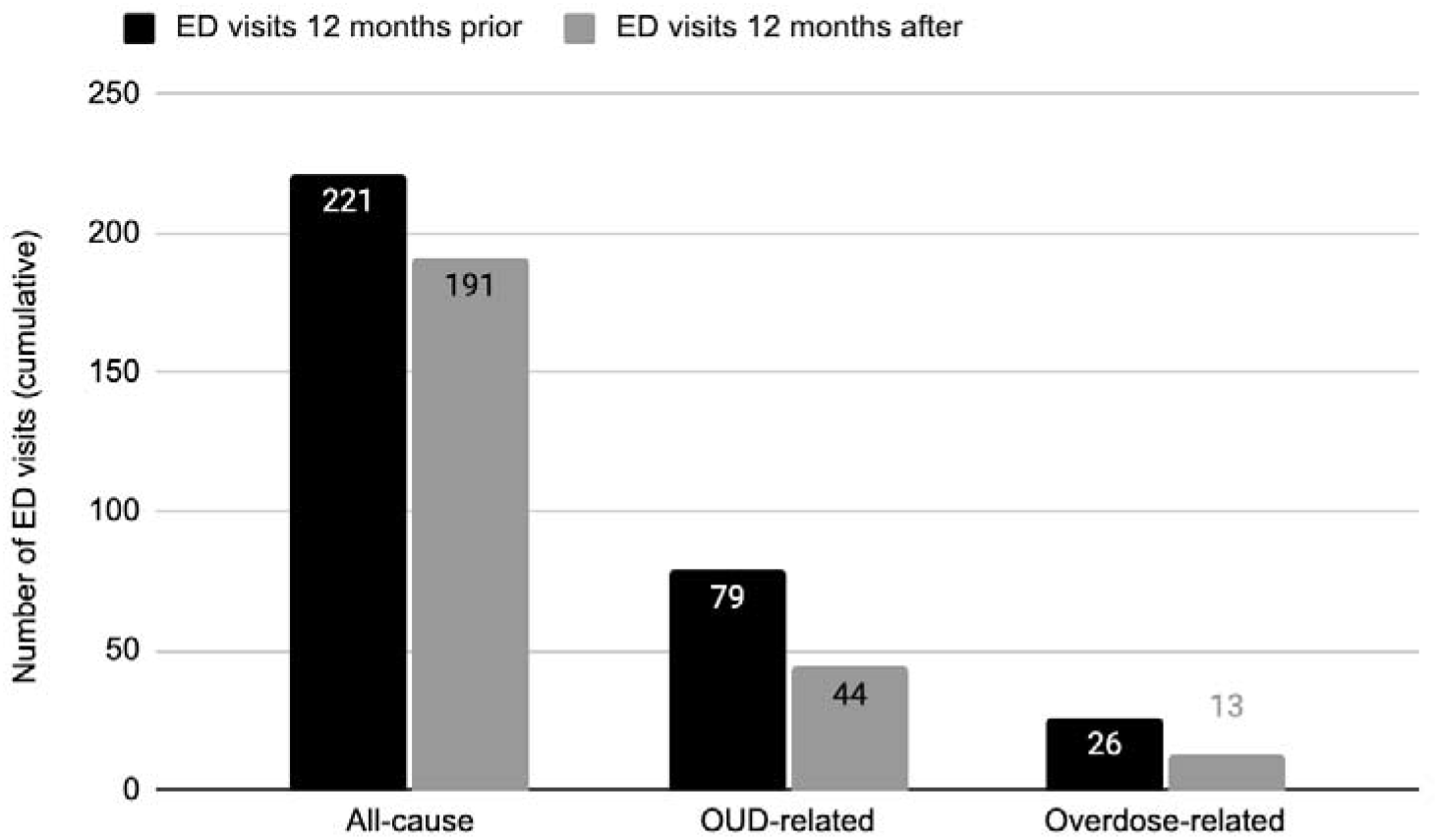
Cohort ED visits 12 months before and after buprenorphine initiation

In the 12 months prior to buprenorphine initiation, the cohort had 79 ED visits related to OUD, overdose, and/or withdrawal. These OUD-related ED visits declined significantly by 44.3% following buprenorphine prescription (IRR = 0.557, 95% CI: 0.317-0.972, p = 0.040). Similarly, individual-level analysis showed a significant decrease in OUD-related visits (p = 0.001) with the interquartile range narrowing from 0-1 to 0-0, indicating more participants without any OUD-related visits (Figure 2). Ultimately, for OUD-related ED visits, we found significant reductions at both individual and cohort levels (Table 2, Figure 1).

**Figure 2.**
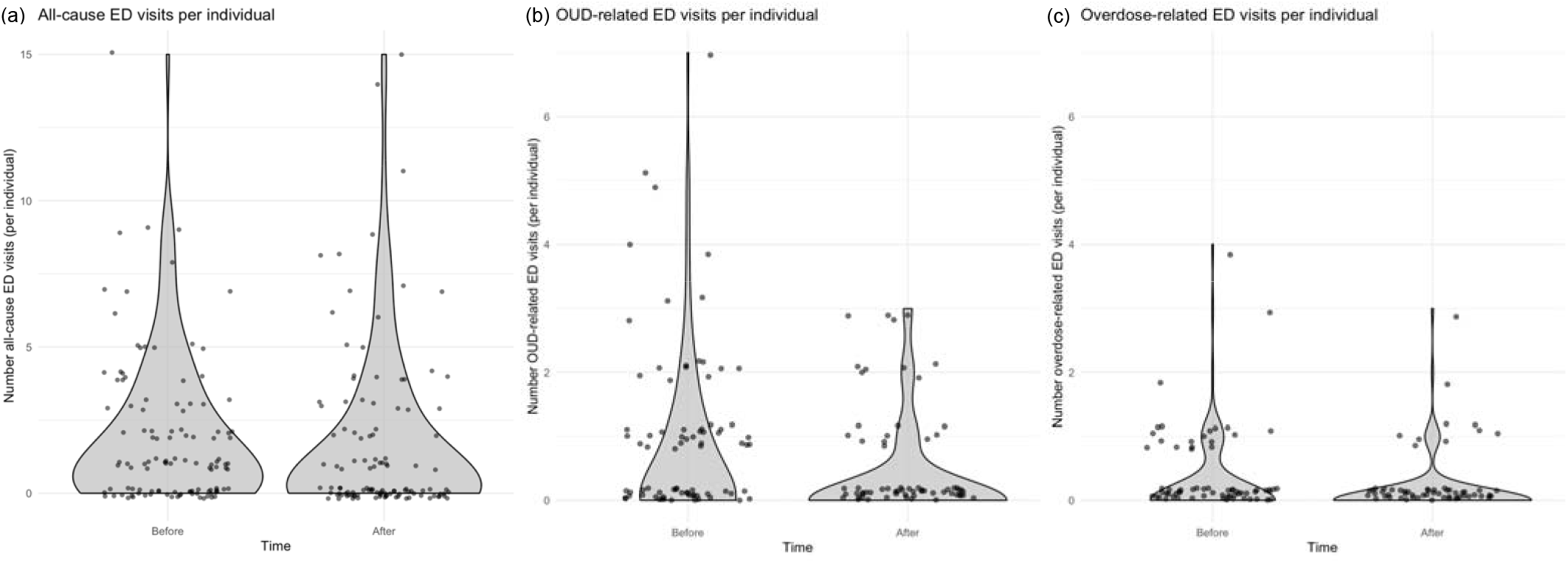
Individual ED visits 12 months before and after buprenorphine

For overdose-specific ED visits, individual-level analysis showed significant reduction after buprenorphine initiation (p = 0.046), though the median and IQR remained at 0 due to the relative rarity of these events (Table 2). At the cohort level, total visits decreased from 26 to 13 (50% reduction), and while this demonstrates a substantial reduction in visit rates (IRR = 0.500, 95% CI: 0.224-1.078, p = 0.082), the trend did not reach significance at the p < 0.05 threshold.

### 3.3 Outliers

Analysis of individual utilization patterns revealed outliers. For all-cause ED visits, one subject increased from 1 visit pre-intervention to 15 visits post-intervention, while two others decreased from 7 visits to 0 visits. For OUD-related ED visits, one subject increased from 2 visits pre-intervention to 10 visits post-intervention, while another decreased from 7 visits to 0 visits. All outliers were included in analysis.

### 3.4 Subgroup analysis

We conducted subgroup analyses to examine whether the intervention’s effectiveness varied by gender, age, and prescription renewal status (Table 3). Subgroup analysis was conducted on change in all-cause ED visits and OUD-related visits. Overdose-related visits were not assessed by subgroup due to the relative rarity of these events.

**Table 3.** Subgroup analysis of change in ED visits.

| **Table 3. Subgroup analysis of change in ED visits** |  |  |  |  |  |
| --- | --- | --- | --- | --- | --- |
| Subgroup | N | Change in all-cause ED visits<sup>*</sup> | P-value<sup>†</sup> | Change in OUD-related ED visits<sup>*</sup> | P-value<sup>†</sup> |
| **Gender** |  |  |  |  |  |
| Male | 68 | 0 (-1.25 to 1) | 0.179 | 0 (0 to 0) | 0.349 |
| Female | 47 | -1 (-2 to 0) |  | 0 (-1 to 0) |  |
| **Age** |  |  |  |  |  |
| Younger | 40 | -1 (-2 to 0) | 0.254 | 0 (-1 to 0) | 0.405 |
| Older | 75 | 0 (-1.5 to 0.5) |  | 0 (-1 to 0) |  |
| **Prescription renewal** |  |  |  |  |  |
| No | 64 | -1 (-2 to 0) | 0.022 | 0 (-1 to 0) | >0.99 |
| Yes | 51 | 0 (-1 to 1.5) |  | 0 (-0.5 to 0) |  |
| <sup>*</sup>Changes in visits are calculated as (visits after – visits before). Negative values indicate fewer visits after the intervention. Values shown are median (IQR).<br><sup>†</sup>Wilcoxon rank-sum test on difference in change in ED utilization between subgroups<br>ED = emergency department; OUD = opioid use disorder |  |  |  |  |  |

Female patients showed a median reduction of 1 visit in all-cause ED utilization (IQR: -2 to 0) while male patients showed no median change (IQR: -1.25 to 1), though this difference was not significant (p = 0.179). Change in OUD-related utilization was also similar between sexes (p = 0.349) with both groups showing a median change of 0 OUD-related visits

Age group analyses comparing younger (age 18-35 years) versus older patients (age 36+ years), found that younger patients had a median reduction of 1 all-cause ED visit (IQR: -2 to 0), while older patients showed no median change (IQR: -1.5 to 0.5), though this difference was not significant (p = 0.254). For OUD-related visits, both age groups showed a median change of 0 (IQR: -1 to 0), with no significant difference between groups (p = 0.405).

Prescription renewal status was not associated with differential changes in OUD-related ED visits, with both groups showing a median change of 0 visits (p > 0.99).

Prescription renewal status was associated with a significant difference in all-cause ED use change. Patients with prescription renewal showed no median change in visits after initiation (IQR: -1 to 1.5) while those without prescription renewal showed a median reduction of 1 visit (IQR: -2 to 0). This difference in change in all-cause ED utilization between subgroups was significant (p = 0.022). Further analysis spurred by this finding showed that among participants with no prescription renewal, the reduction in all-cause ED visits after buprenorphine initiation compared to before was significant (Wilcoxon signed rank test p=0.003). Among participants with renewed prescriptions there was no significant change in all-cause ED visits after buprenorphine initiation (Wilcoxon signed rank test p = 0.648).

## 4. Discussion

Initiation of buprenorphine via the SM mobile unit was associated with significantly decreased incidence of OUD-related ED visits at both cohort and individual levels. Buprenorphine initiation was also associated with a decrease in overdose-specific ED visits at an individual level. There was a trend toward decreased overdose-specific ED visits at the cohort level that did not reach significance, likely due to the relatively low frequency of these events. These results suggest that SM-initiated buprenorphine is a successful modality for reducing addiction-related healthcare utilization among PEH. This aligns with other investigations of decentralized, low-barrier MOUD initiation that have shown positive outcomes, such as higher rates of patient retention in treatment^21^ and patient satisfaction^22^ compared to patients who utilized fixed-site services.^23^

In our study, all-cause ED visits did not decrease significantly after buprenorphine initiation. PEH face wide-ranging health challenges, with evidence suggesting that the majority of PEH with substance use disorder have co-occurring physical or mental illness.^24^ The significant burden of chronic health concerns besides addiction may explain why overall ED visits did not significantly change at a cohort or individual level. The decrease in OUD-related visits compared to steady all-cause ED utilization points to individuals in this study continuing to seek care for concerns unrelated to OUD after buprenorphine initiation, even as their OUD-related needs decreased. While increased healthcare utilization is commonly interpreted as a marker of greater healthcare need and worse health, this picture is problematized in the setting of severe barriers to care where increased utilization can indicate improved access rather than increased need.

Other studies have also investigated the relationship between low barrier MOUD for PEH and healthcare utilization. While MOUD has been associated with significantly reduced ED and hospital utilization among people who use drugs,^25^ Fine et al found that patients who had an encounter with an addiction-focused mobile health unit had a significant increase in both hospitalizations and ED utilization in the subsequent year compared with up to four years prior.^26^ This discrepancy may be due in part to the fact that patients reached through mobile health units for addiction treatment (as in Fine et al’s population), are more likely to be homeless, have more severe addiction and mental illness, and inject drugs, all of which may lead to greater healthcare need and utilization.^23,27,28^

Among patients in our study initiated on buprenorphine by SM, 30% renewed their prescriptions with a non-SM provider, 14% renewed prescriptions with SM, and 55% did not renew their prescription. While return to use is a possible reason for non-renewal, there are many others including mortality, incarceration, moving out of state (where prescription renewal would not be in the study state’s PDMP), initiation on a different form of MOUD (e.g. methadone), or entry to an addiction treatment program that doesn’t a) allow buprenorphine or b) enter buprenorphine data into PDMP. Through provider knowledge of patient history, we know that at least four patients started residential treatment 5-12 months after buprenorphine prescription, one patient who did not renew their prescription re-started buprenorphine 12 months after initial prescription.

Patients with no prescription renewal had a significant reduction in all-cause ED utilization, in contrast to patients who did renew their prescription, for whom there was no significant change. This is an interesting finding in the setting of insignificant change in all-cause ED utilization overall, and significantly reduced OUD-related ED use among both groups. This may have occurred because patients who renewed their prescription maintained greater connection with local healthcare systems and were therefore relatively more likely to seek care for non-OUD related medical concerns than patients who did not renew their prescription. Alternatively, the decrease in all-cause ED use among non-renewers could be driven by factors that would lead to both lack of prescription renewal (as measured by the study state’s PDMP) and reduction in all-cause ED use (as measured by the study hospital system’s EMR, which receives only in-state ED visit records) including reasons for non-renewal discussed above, like moving out of state or methadone initiation. Etiologies like moving out of state would reduce all types of ED use, while we would hypothesize that initiating alternative addiction treatment would reduce OUD-related ED use but not necessarily all-cause ED use. Ultimately, without greater detail about reasons for non-renewal, it is difficult to draw conclusions about the decrease in all-cause ED use among non-renewers relative to those who renewed their prescription.

As treatment for opioid use disorder has expanded, buprenorphine uptake has increased inequitably across the US, with increases in buprenorphine prescriptions and providers occurring in predominantly White, wealthier neighborhoods.^29,30^ Interventions like ED Peer Navigator Programs expand MOUD prescription and linkage to addiction care to people who may not have an office-based primary care provider, and have been shown to decrease ED use and hospitalization.^31^ Our study similarly demonstrates that the expansion of accessible and consistent MOUD for patients who are chronically homeless and who have not been the primary recipients of this evidence-based treatment is associated with positive outcomes. These findings should build upon the evidence base that highlights the need to expand buprenorphine to patients who otherwise may be systematically excluded from accessing and maintaining treatment.

### 4.1 Future research

Given that our study showed no change in all-cause ED visits but a decline in OUD-related utilization after MOUD initiation, this may indicate consistent access to care alongside less OUD-associated morbidity. Further studies correlating healthcare utilization, reasons for utilization, and markers of overall health are needed to elucidate the complex relationship between mobile health units, healthcare utilization, and patient wellbeing. We also propose that future research should investigate ED utilization for non-OUD related chief complaints among patients who renew vs. do not renew buprenorphine prescription, to understand the driving force behind the decrease in all-cause ED use among non-renewers. Studies that assess more direct indicators of patient health than healthcare utilization, such as mortality or illness-free days, could further elucidate the relationship between SM-initiated MOUD, healthcare utilization, and patient health. Further understanding reasons for prescription renewal with the SM program versus other providers, as well as reasons for lack of prescription renewal, may also help inform development of sustainable MOUD treatment models for PEH. Collecting meaningful qualitative data may also provide valuable insights into the barriers and facilitators that influence follow-up adherence among PEH receiving MOUD. Finally, studying the cost effectiveness of reductions in ED utilization can help demonstrate the overall value of mobile health units.

### 4.2 Limitations

While providing strong evidence for the impact of buprenorphine initiation on ED utilization, this analysis should be considered in light of its limitations in study design, outcomes measures, and dataset constraints. The present study is limited by the fact that it does not include outcome measures that are common across other studies—such as retention data on MOUD at regular time points—which makes it less comparable to other literature published on mobile health units providing MOUD.^21,32–35^ Unlike Hall et al. and Fine et al., there is no comparison group to patients who initiated and maintained treatment on MOUD via traditional means at a fixed-location prescriber, which limits our ability to speak to the efficacy of this intervention.^26,27^ Further, this analysis does not assess other forms of healthcare utilization, such as hospitalization or office visits, which may also be influenced by MOUD initiation or contact with a mobile health unit. Moreover, ED utilization, while an accessible proxy for both morbidity and healthcare costs, is far from a perfect measure of either patients’ wellbeing or cost of care.

Finally, this analysis only considers age and sex demographic data and does not consider other important demographic data like race and ethnicity. Pinkhover et al. highlights that racial and ethnic minorities have been disproportionately impacted by existing failures in MOUD treatment, including lower likelihood of receiving buprenorphine for MOUD compared to White patients, and an increased likelihood of receiving ineffective doses and durations of treatment.^29^ These patients also experience greater loss to follow up compared to their White counterparts. Mobile health units delivering MOUD have the potential to better serve low-income communities of color, and it is essential to center health equity in these interventions to prevent perpetuating inequities associated with this life-saving treatment.

## 5. Conclusions

Treating addiction and preventing overdose in PEH remains a pressing public health emergency. This study observed that buprenorphine initiation by a SM program was associated with a significant decline in OUD-related ED visits at both cohort and individual levels, and a notable reduction in overdose-related visits. Furthermore, a large portion of patients continued their buprenorphine prescription. Taken together, these findings support SM-initiated buprenorphine as a strategy for reducing opioid-related ED utilization while encouraging sustained engagement in care. SM programs represent promising avenues to initiate buprenorphine and reduce OUD-related morbidity among people experiencing homelessness.

## Data Availability

All data produced in the present study are available upon reasonable request to the authors

## Acknowledgements

The authors thank the Mt. Carmel Street Medicine Program team for their dedication, example, and support, and The Ohio State Emergency Department for supporting submission fees. This research did not receive any specific grant from funding agencies in the public, commercial, or not-for-profit sectors.

## Notes

### Competing Interest Statement

The authors have declared no competing interest.

### Funding Statement

This study did not receive any funding

### Author Declarations

The IRB of Mount Carmel Health System gave ethical approval for this work

